# Vascular risk burden is a key player in the early progression of Alzheimer’s disease

**DOI:** 10.1101/2021.12.18.21267994

**Authors:** João Pedro Ferrari-Souza, Wagner S. Brum, Lucas A. Hauschild, Lucas U. Da Ros, Pâmela C. Lukasewicz Ferreira, Bruna Bellaver, Douglas T. Leffa, Andrei Bieger, Cécile Tissot, Marco Antônio De Bastiani, Guilherme Povala, Andréa L. Benedet, Joseph Therriault, Yi-Ting Wang, Nicholas J. Ashton, Henrik Zetterberg, Kaj Blennow, Sheila O. Martins, Diogo O. Souza, Pedro Rosa-Neto, Thomas Karikari, Tharick A. Pascoal, Eduardo R. Zimmer, for the Alzheimer’s Disease Neuroimaging Initiative

## Abstract

Understanding whether vascular risk factors synergistically potentiate Alzheimer’s disease progression is important in the context of emerging treatments for preclinical Alzheimer’s disease. The existence of a synergistic relationship could suggest that the combination of therapies targeting Alzheimer’s disease pathophysiology and vascular risk factors might potentiate treatment outcomes. In the present retrospective cohort study, we tested whether vascular risk factor burden interacts with Alzheimer’s disease pathophysiology to accelerate neurodegeneration and cognitive decline in cognitively unimpaired subjects. We evaluated 503 cognitively unimpaired participants from the Alzheimer’s Disease Neuroimaging Initiative (ADNI) study. Baseline vascular risk factor burden was calculated considering the history of cardiovascular disease, hypertension, diabetes mellitus, hyperlipidemia, stroke or transient ischemic attack, smoking, atrial fibrillation, and left ventricular hypertrophy. Alzheimer’s disease pathophysiology was evaluated using cerebrospinal fluid (CSF) amyloid-β_1-42_ (Aβ_1-42_) reflecting brain amyloidosis (A) and tau phosphorylated at threonine 181 (p-tau_181_) reflecting brain tau pathology (T). Individuals were dichotomized as having an elevated vascular risk factor burden (V+ if having two or more vascular risk factors) and as presenting preclinical Alzheimer’s disease [(AT)+ if having abnormal CSF p-tau_181_ and Aβ_1-42_ levels]. Neurodegeneration was assessed with plasma neurofilament light (NfL) and global cognition with the modified version of the Preclinical Alzheimer’s Cognitive Composite. Linear mixed-effects models revealed that an elevated vascular risk factor burden synergistically interacted with Alzheimer’s disease pathophysiology to drive longitudinal increases in plasma NfL levels (β = 5.08, *P* = 0.016) and cognitive decline (β = −0.43, *P* = 0.020). Additionally, we observed that vascular risk factor burden was not associated with CSF Aβ_1-42_ or p-tau_181_ changes over time. Survival analysis demonstrated that individuals with preclinical Alzheimer’s disease and elevated vascular risk factor burden [(AT)+V+] had a significantly greater risk of clinical progression to cognitive impairment (adjusted Hazard Ratio = 3.5, *P* < 0.001). Our results support the notion that vascular risk factor burden and Alzheimer’s disease pathophysiology are independent processes; however, they synergistically lead to neurodegeneration and cognitive decline. These findings can help in providing the blueprints for the combination of vascular risk factor management and Alzheimer’s disease pathophysiology treatment in preclinical stages. Moreover, we observed plasma NfL as a robust marker of disease progression that may be used to track therapeutic response in future trials.

## 1. Introduction

A new era of trials in individuals with preclinical Alzheimer’s disease is starting given the assumption that better outcomes could be achieved with interventions performed before the presence of extensive damage and cognitive symptoms.^1, 2^ The preclinical stage of Alzheimer’s disease has been characterized as the presence of biomarker evidence of amyloid-β (Aβ) and tau pathologies in cognitively unimpaired (CU) individuals.^3^ Subjects in this stage are at higher risk for Alzheimer’s disease clinical progression; however, many of them never progress to cognitive impairment, suggesting that other simultaneous pathological processes are involved.^4^ Therefore, it is important to understand the additional factors contributing to Alzheimer’s disease progression for the development of effective therapeutic strategies.

Vascular risk factors (VRFs), such as hypertension, diabetes mellitus, smoking, and hypercholesterolemia, are well-established risk factors for developing Alzheimer’s disease dementia.^5–11^ These conditions are associated with cerebrovascular lesions in neuropathologically-confirmed Alzheimer’s disease patients,^12, 13^ and the presence of these brain injuries contributes to dementia onset.^12–15^ Furthermore, as multiple VRFs often coexist and gradually increase Alzheimer’s disease risk,^6^ recent research focused on these conditions in combination (i.e., burden) rather than individually. Nevertheless, it remains to be elucidated whether vascular risk and Alzheimer’s disease pathophysiology have additive or synergistic effects on neurodegeneration and cognitive decline. Also, the direct effect of VRFs on Alzheimer’s disease pathophysiology is still not completely understood. While some studies support a possible relation with Aβ or tau deposition,^16–19^ others point to the opposite direction.^12, 20–23^

Determining whether vascular risk factors synergistically potentiate Alzheimer’s disease progression has potential implications for clinical trials. It could suggest that the combination of therapies targeting Aβ and tau pathologies, as well as VRFs, may enhance treatment outcomes. With this in mind, the main goal of the present study was to investigate whether VRF burden synergistically interacts with Alzheimer’s disease pathophysiology to accelerate neurodegeneration and cognitive decline in CU individuals. Secondarily, we also aimed to assess whether VRF burden is related to changes in Aβ and tau biomarkers over time.

## 2. Materials and methods

Data used in the preparation of this article were obtained from the Alzheimer’s Disease Neuroimaging Initiative (ADNI) database (adni.loni.usc.edu; last accessed May 2021). The ADNI was launched in 2003 as a public-private partnership, led by Principal Investigator Michael W. Weiner, MD. The primary goal of ADNI has been to test whether serial magnetic resonance imaging (MRI), positron emission tomography (PET), other biological markers, and clinical and neuropsychological assessment can be combined to measure the progression of mild cognitive impairment (MCI) and early Alzheimer’s disease. Detailed information concerning inclusion and exclusion criteria has already been described.^24^ Of note, participants were recruited between the ages of 55 and 90 years, completed at least 6 years of education, were fluent in Spanish or English, had a Hachinski ischemic score less than or equal to four, and had screening/baseline MRI scans without evidence of infection, infarction, or other focal lesions (subjects with multiple lacunes or lacunes in a critical memory structure were excluded). Institutional Review Boards of all involved sites approved the ADNI study and all research participants or their authorized representatives provided written informed consent.

### 2.1. Participants

We evaluated CU individuals from the ADNI cohort. All participants presented Mini-Mental State Examination (MMSE) scores ≥ 24 and Clinical Dementia Rating (CDR) of 0. Subjects did not have any significant neurological disease. To investigate the longitudinal cognitive trajectory and risk of clinical progression to cognitive impairment, we assessed 503 individuals with available baseline medical data and cerebrospinal fluid (CSF) Elecsys biomarkers (Aβ_1-42_ and p-tau_181_), as well as longitudinal clinical assessments with neuropsychological testing (up to 6 years). We restricted these analyses to subjects with CSF collected within 1.2 years of the first neuropsychological assessment.

Analyses evaluating the longitudinal trajectories of fluid biomarkers were performed in subsamples based on specific data availability. To assess neurodegeneration over time, individuals with longitudinal plasma NfL measurements (up to 4 years) were included (*n* = 269). To assess changes in Alzheimer’s disease CSF biomarkers, individuals with longitudinal CSF Aβ_1-42_ and p-tau_181_ measurements (up to 6 years) were included (*n* = 284). More details regarding patient selection criteria are provided in **Supplementary Methods 1**.

### 2.2. VRF burden

Information regarding medical history and use of medications was manually assessed in ADNI records by three independent investigators (JPFS, LAH, and LUDR) to determine VRF burden. A previously proposed composite score to estimate the lifetime risk of cardiovascular disease^25, 26^ was adapted to assess cerebrovascular injuries in Alzheimer’s disease patients.^12, 13^ Baseline VRF burden was calculated using the modified score,^12, 13^ which considers the presence or absence of history for the following conditions: (i) cardiovascular disease [coronary artery disease (myocardial infarction, angina, stent placement, angioplasty, coronary artery bypass graft, coronary insufficiency), heart failure, or intermittent claudication]; (ii) hypertension (positive medical history or use of antihypertensive medications), (iii) diabetes mellitus (positive medical history or use of antidiabetic therapy), (iv) hyperlipidemia (positive medical history or use of lipid-lowering drugs); (v) stroke or transient ischemic attack (TIA); (vi) smoking (ever or never); (vii) atrial fibrillation; and (viii) left ventricular hypertrophy. Since total burden was calculated by the sum of VRFs, each participant presented a composite score from zero to eight. Noteworthy, the score used does not have different weights for individual VRFs. Further information regarding VRF burden assessment can be found in **Supplementary Methods 2**. Flowchart of medication assessment is shown in **Supplementary Fig. 1**. A detailed list of all included drugs detected in ADNI records for hypertension, diabetes mellitus, and hyperlipidemia is reported in **Supplementary Table 1**.

### 2.3. Biomarkers

CSF Aβ_1-42_ and p-tau_181_ levels were measured using fully automated Elecsys immunoassays (Roche Diagnostics).^27, 28^ Measurements outside the analytical range (< 200 pg/mL or > 1700 pg/mL for Aβ_1-42_; < 8 pg/mL or > 120 pg/mL for p-tau_181_) were set to their respective technical limit. Of note, we did not use CSF total tau (t-tau) due to its strong correlation with CSF p-tau_181_.^29^ Plasma NfL levels were analyzed using an in-house immunoassay on the Single molecule array (Simoa) platform (Quanterix Corporation).^30, 31^ Three subjects presenting NfL concentrations three standard deviations (SD) above or below the mean of the whole population at baseline were considered outliers and, thus, excluded from the analysis assessing plasma NfL trajectory. To directly assess cerebrovascular disease in exploratory analyses, we quantified white matter hyperintensity (WMH) volumes using previously described automated methods.^32, 33^

### 2.4. Cognition

The modified version of Preclinical Alzheimer’s Cognitive Composite (mPACC)^34, 35^ was used as an outcome to evaluate the cognitive trajectory of participants as it was developed to detect cognitive changes in CU individuals with biomarker evidence of Alzheimer’s disease pathophysiology and adapted for the ADNI study. The mPACC was calculated by averaging the z-score of the following tests: MMSE, delayed recall for the Alzheimer’s Disease Assessment Scale – Cognitive Subscale (ADAS-Cog), Logical Memory Delayed Recall, and the Trail Making Test B. To evaluate clinical progression to cognitive impairment, a clinical diagnosis of MCI or dementia in follow-up visits – accordingly to ADNI protocols - was used as an outcome. Moreover, we also used the clinical dementia rating scale sum of boxes (CDR-SB) to assess clinical deterioration in exploratory analyses.

### 2.5. Cutpoints

Aβ positivity was defined as CSF Aβ_1-42_ < 977 pg/mL and p-tau positivity was determined as CSF p-tau_181_ > 24 pg/mL.^36, 37^ Preclinical Alzheimer’s disease [(AT)+] was defined as positivity for both biomarkers simultaneously (i.e., A+T+). Other groups (i.e., A+T-, A-T+ or A-T-) were considered as not having preclinical Alzheimer’s disease [(AT)-].^3^

Neuropathologically-confirmed Alzheimer’s disease patients with two or more of the VRFs investigated are more likely to present occult cerebrovascular changes at autopsy; however, the presence of just one VRF is not necessarily associated with brain vascular lesions in these patients.^12, 13^ Therefore, an elevated VRF burden (V+) was defined as a vascular composite score equal or higher than two; individuals with composite score equal to zero or one were classified as having a low VRF burden (V-). This threshold is especially applicable for the ADNI cohort since the presence of significant cerebrovascular lesions is an exclusion criterion at study enrollment. In the exploratory analyses, we divided participants into low and high WMH groups (WMH- and WMH+, respectively) based on median split; thresholds were calculated separately in each method used to quantify WMH volume.

### 2.6. Statistical analysis

We used the R Statistical Software (version 4.0.2, http://www.r-project.org/) to perform all statistical analyses. Comparison between the groups’ demographic characteristics was done with descriptive statistics. Linear mixed-effects (LME)-based analyses and survival analyses were carried out using the “lme4” and “survival” packages, respectively.

For all analyses, VRF burden status refers to a dichotomous variable (V- *vs* V+), as well as Alzheimer’s disease pathophysiology status [(AT)- *vs* (AT)+]. Moreover, baseline was defined as the first outcome measurement for each analysis. Statistical significance level was set as P < 0.05, two-tailed.

#### 2.6.1. LME-based analysis

At first, LME models were performed to evaluate the existence of a synergistic relationship between VRF burden and Alzheimer’s disease pathophysiology, as well as their independent effects, on plasma NfL levels (Model A) and cognitive performance (Model B). In each model, besides evaluating VRF burden status and Alzheimer’s disease pathophysiology status main effects, we also assessed the two-way interactions of VRF burden status with time, Alzheimer’s disease pathophysiology status with time, and VRF burden status with Alzheimer’s disease pathophysiology status, as well as the three-way interaction of VRF burden status and Alzheimer’s disease pathophysiology status with time. For visualization purposes, graphs were plotted stratifying participants into four groups according to VRF burden status and Alzheimer’s disease pathophysiology status [(AT)-V-, (AT)-V+, (AT)+V-, and (AT)+V+]. To confirm the presence of a synergistic relationship, we tested whether the interaction effects of VRF burden and Alzheimer’s disease pathophysiology were greater than the sum of their independent effects.^38–40^ Here we were mainly interested in the rates of changes in plasma NfL levels and mPACC scores. Therefore, the independent effects correspond to the absolute β coefficients of the two-way interactions of VRF burden with time and Alzheimer’s disease pathophysiology with time; the interaction effects correspond to the absolute β coefficients of the three-way interactions of VRF burden status and Alzheimer’s disease pathophysiology status with time.

Secondarily, we also used LME models to determine the association of VRF burden status with changes in CSF Aβ_1-42_ (Model C) and p-tau_181_ levels (Model D) over time. In these analyses, both CSF Aβ_1-42_ or p-tau_181_ were treated as continuous variables and we were mainly interested in the effects of VRF burden and its interaction with time.

LME models were fit including subject-specific random slopes and intercepts. All models were adjusted for age, sex, years of education, apolipoprotein E ε4 (*APOE* ε4) status, and their interactions with time,^41^ to properly account for potential confounders and to avoid significance-based selection.^42^ Furthermore, continuous predictors were standardized, and time was treated as a continuous variable (years from baseline). Confidence intervals were estimated using bootstrapping with 1000 permutations, and the presence of multicollinearity was appraised by calculating the variance inflation factors (VIFs).

#### 2.6.2. Survival analysis

Time-to-event analysis was carried to evaluate the risk of clinical progression to cognitive impairment (i.e., MCI or dementia) according to VRF burden and Alzheimer’s disease pathophysiology status. Participants contributed to follow-up time from first clinical and neuropsychological assessment until clinical conversion (i.e., CU-to-MCI or CU-to-dementia), loss to follow-up, or end of follow-up (6 years after baseline). Kaplan-Meier curves display observed survival probabilities across the four groups [(AT)-V-, (AT)-V+, (AT)+V-, and (AT)+V+], which was statistically compared using the generalized log-rank test. Adjusted hazard ratios (HRs) were calculated using a Cox-proportional hazards model that was fitted with the following predictors: group, age, sex, years of education, and *APOE* ε4 status. Post-hoc pairwise contrast analyses with Tukey’s multiple comparison test were performed to compare the adjusted HR among groups. As sensitivity analysis, we performed an additional Cox-proportional hazards model only fitting group as the predictor to determine crude HRs (i.e., unadjusted estimates). Concerning model diagnostics, proportional hazards assumption was assessed by Schoenfeld residuals. Similar to LME models, continuous predictors were standardized and time was handled as a continuous variable (years from baseline). To better understand the risk of clinical progression across groups, we conducted exploratory analyses (i) using changes in the CDR-SB as outcome, (ii) restricting to individuals under 75 years old, and (iii) using WMH status instead of VRF burden status to divide the groups.

## 3. Results

A total of 503 participants (43.7% males) were assessed in this study, of whom 13.1% had CSF biomarker evidence of preclinical Alzheimer’s disease [(AT)+], and 45.9% presented an elevated VRF burden (V+). Sample demographics, biomarker and clinical characteristics are summarized in **Table 1**. In addition, detailed demographic information regarding subsamples used to evaluate the trajectory of plasma NfL and CSF biomarkers - Aβ_1-42_ and p-tau_181_ – are available in the **Supplementary Table 2** and **Supplementary Table 3**. Regarding model inspections, multicollinearity was not identified in any LME model used. Besides, survival model diagnostics indicated that proportional hazards assumption was not violated.

**Table 1.** Demographics and key characteristics of participants^a^.

|  | (AT)-V- | (AT)-V+ | (AT)+V- | (AT)+V+ |
| --- | --- | --- | --- | --- |
| No. | 238 | 199 | 34 | 32 |
| Age at baseline, y | 72.2 (6.3) | 73.2 (6.0) | 72.9 (5.9) | 77.4 (5.0) |
| Male, No. (%) | 83 (34.9) | 105 (52.8) | 16 (47.1) | 16 (50.0) |
| Education, y | 16.6 (2.6) | 16.5 (2.6) | 16.9 (2.5) | 15.8 (2.2) |
| *APOE* ε4 carriers, No. (%) | 57 (23.9) | 54 (27.1) | 22 (64.7) | 18 (56.3) |
| Individual VRFs at baseline, No. (%)<sup>b</sup> |  |  |  |  |
| Cardiovascular disease | 2 (0.8) | 31 (15.6) | 0 (0.0) | 8 (25.0) |
| Hypertension | 48 (20.2) | 169 (84.9) | 7 (20.6) | 27 (84.4) |
| Diabetes mellitus | 1 (0.4) | 40 (20.1) | 0 (0.0) | 4 (12.5) |
| Atrial fibrillation | 1 (0.4) | 10 (5.0) | 0 (0.0) | 3 (9.4) |
| Smoking | 27 (11.3) | 65 (32.7) | 4 (11.8) | 11 (34.4) |
| TIA / stroke | 1 (0.4) | 12 (6.0) | 0 (0.0) | 2 (6.3) |
| Hyperlipidemia | 58 (24.4) | 173 (86.9) | 11 (32.4) | 30 (93.8) |
| VRF burden at baseline | 0.6 (0.5) | 2.5 (0.7) | 0.6 (0.5) | 2.7 (0.5) |
| CSF Aβ<sub>1-42</sub> at baseline, pg/mL | 1265.1 (417.1) | 1282.0 (406.7) | 691.5 (165.0) | 703.6 (177.9) |
| CSF p-tau<sub>181</sub> at baseline, pg/mL | 19.1 (7.4) | 20.1 (7.6) | 33.5 (7.2) | 35.6 (10.8) |
| Plasma NfL at baseline, pg/mL<sup>c</sup> | 32.3 (13.6) | 33.0 (14.5) | 35.6 (5.5) | 41.2 (10.5) |
| MMSE score at baseline | 29.2 (1.1) | 29.0 (1.2) | 29.3 (0.9) | 28.9 (1.3) |
| mPACC score at baseline | 0.5 (2.4) | -0.3 (2.6) | -0.1 (2.5) | -1.7 (2.4) |
| Follow-up, y | 3.6 (1.8) | 3.8 (1.8) | 3.5 (1.9) | 3.7 (1.8) |

Participants were stratified according to vascular risk factor (VRF) burden status and Alzheimer's disease pathology status. Continuous variables are presented as mean (SD). *APOE* ε4 = Apolipoprotein E ε4; Aβ<sub>1-42</sub> = amyloid-β<sub>1-42</sub>; CSF = cerebrospinal fluid; MMSE = Mini-Mental State Examination; mPACC = modified Preclinical Alzheimer's Cognitive Composite; NfL = neurofilament light; p-tau<sub>181</sub> = tau phosphorylated at threonine 181; TIA = transient ischemic attack.
<sup>a</sup>In this table, baseline refers to the visit of first clinical assessment with neuropsychological testing.
<sup>b</sup>Prevalence of left ventricular hypertrophy is not displayed in the table because only one participant in the (AT)-V+ group was described to have this condition in the Alzheimer's Disease Neuroimaging Initiative database.
<sup>c</sup>Assessed in a subset of 229 subjects who had available plasma NfL measurement at the same visit of first neuropsychological assessment.

### 3.1. VRF burden and Alzheimer’s disease pathophysiology act synergistically on plasma NfL levels

LME model coefficients for the associations between VRF burden, Alzheimer’s disease pathophysiology, and longitudinal plasma NfL can be found in **Table 2**, Model A. At baseline, preclinical Alzheimer’s disease status was marginally associated with higher concentrations of plasma NfL (β = 6.23, *P* = 0.082). On the other hand, VRF burden was not significantly associated with baseline plasma NfL levels (*P* = 0.542), nor was the interaction between VRF burden and Alzheimer’s disease pathophysiology (*P* = 0.754). Concerning plasma NfL longitudinal trajectory, there was a significant three-way interaction (VRF burden x Alzheimer’s disease pathophysiology x time; β = 5.08, *P* = 0.016), indicating that an elevated VRF burden acted synergistically with preclinical Alzheimer’s disease to increase plasma NfL concentrations longitudinally. To confirm the presence of a synergistic interaction, we found that the interaction effect of VRF burden and Alzheimer’s disease pathophysiology was greater than the sum of their independent effects (**Table 3**, Model A). Interestingly, VRF burden x time and Alzheimer’s disease pathophysiology x time interaction terms were not significant (*P* = 0.351 and *P* = 0.793, respectively). This shows that VRF burden and Alzheimer’s disease pathophysiology did not associate independently with changes in NfL levels over time. For results stratified by groups, see **Fig. 1A**.

**Table 2.** Liner mixed-effects model coefficients.

| | $\beta$ (95% CI) | T-value | P-value |
| --- | --- | --- | --- |
| **Model A<sup>a</sup>: plasma NfL ~ VRF burden status x Alzheimer's disease pathophysiology status x time + covariates<sup>e</sup> x time** |  |  |  |
| Elevated VRF burden | 1.04 (-2.23 to 4.46) | 0.61 | 0.542 |
| Preclinical Alzheimer's disease | 6.23 (-0.61 to 12.81) | 1.75 | 0.082 |
| Elevated VRF burden x preclinical Alzheimer's disease | -1.44 (-10.75 to 8.39) | -0.31 | 0.754 |
| Elevated VRF burden x time | -0.72 (-2.17 to 0.64) | -0.94 | 0.351 |
| Preclinical Alzheimer's disease x time | 0.42 (-2.73 to 3.45) | 0.26 | 0.793 |
| Elevated VRF burden x preclinical Alzheimer's disease x time | 5.08 (1.41 to 9.23) | 2.43 | 0.016 |
| **Model B<sup>b</sup>: mPACC ~ VRF burden status x Alzheimer's disease pathophysiology status x time + covariates<sup>e</sup> x time** |  |  |  |
| Elevated VRF burden | -0.39 (-0.79 to 0.04) | -1.86 | 0.063 |
| Preclinical Alzheimer's disease | -0.02 (-0.80 to 0.80) | -0.04 | 0.969 |
| Elevated VRF burden x preclinical Alzheimer's disease | -0.58 (-1.77 to 0.49) | -1.01 | 0.312 |
| Elevated VRF burden x time | 0.05 (-0.07 to 0.18) | 0.79 | 0.431 |
| Preclinical Alzheimer's disease x time | -0.22 (-0.49 to 0.04) | -1.67 | 0.096 |
| Elevated VRF burden x preclinical Alzheimer's disease x time | -0.43 (-0.79 to -0.08) | -2.34 | 0.020 |
| **Model C<sup>c</sup>: CSF Aβ<sub>1-42</sub> ~ VRF burden status x time + covariates<sup>e</sup> x time** |  |  |  |
| Elevated VRF burden | -44.33 (-130.95 to 53.20) | -0.90 | 0.367 |
| Elevated VRF burden x time | 1.76 (-11.22 to 14.98) | 0.27 | 0.789 |
| **Model D<sup>d</sup>: CSF p-tau<sub>181</sub> ~ VRF burden status x time + covariates<sup>e</sup> x time** |  |  |  |
| Elevated VRF burden | 0.21 (-2.06 to 2.39) | 0.19 | 0.848 |
| Elevated VRF burden x time | -0.13 (-0.37 to 0.13) | -1.01 | 0.314 |

*APOE* ε4 = Apolipoprotein E ε4; Aβ <sub>1-42</sub> = amyloid-β <sub>1-42</sub>; CI = confidence interval; CSF = cerebrospinal fluid; mPACC = modified Preclinical Alzheimer's Cognitive Composite; NfL = neurofilament light; p-tau<sub>181</sub> = tau phosphorylated at threonine 181; VRF = vascular risk factor.
<sup>a</sup>Marginal $R^2$ : 0.32; Conditional $R^2$ : 0.79.
<sup>b</sup>Marginal $R^2$ : 0.21; Conditional $R^2$ : 0.74.
<sup>c</sup>Marginal $R^2$ : 0.15; Conditional $R^2$ : 0.92.
<sup>d</sup>Marginal $R^2$ : 0.10; Conditional $R^2$ : 0.98.
<sup>e</sup>Potential confounders included in the models as covariates are the following: age, sex, years of education, and *APOE* ε4 status.

**Table 3.**
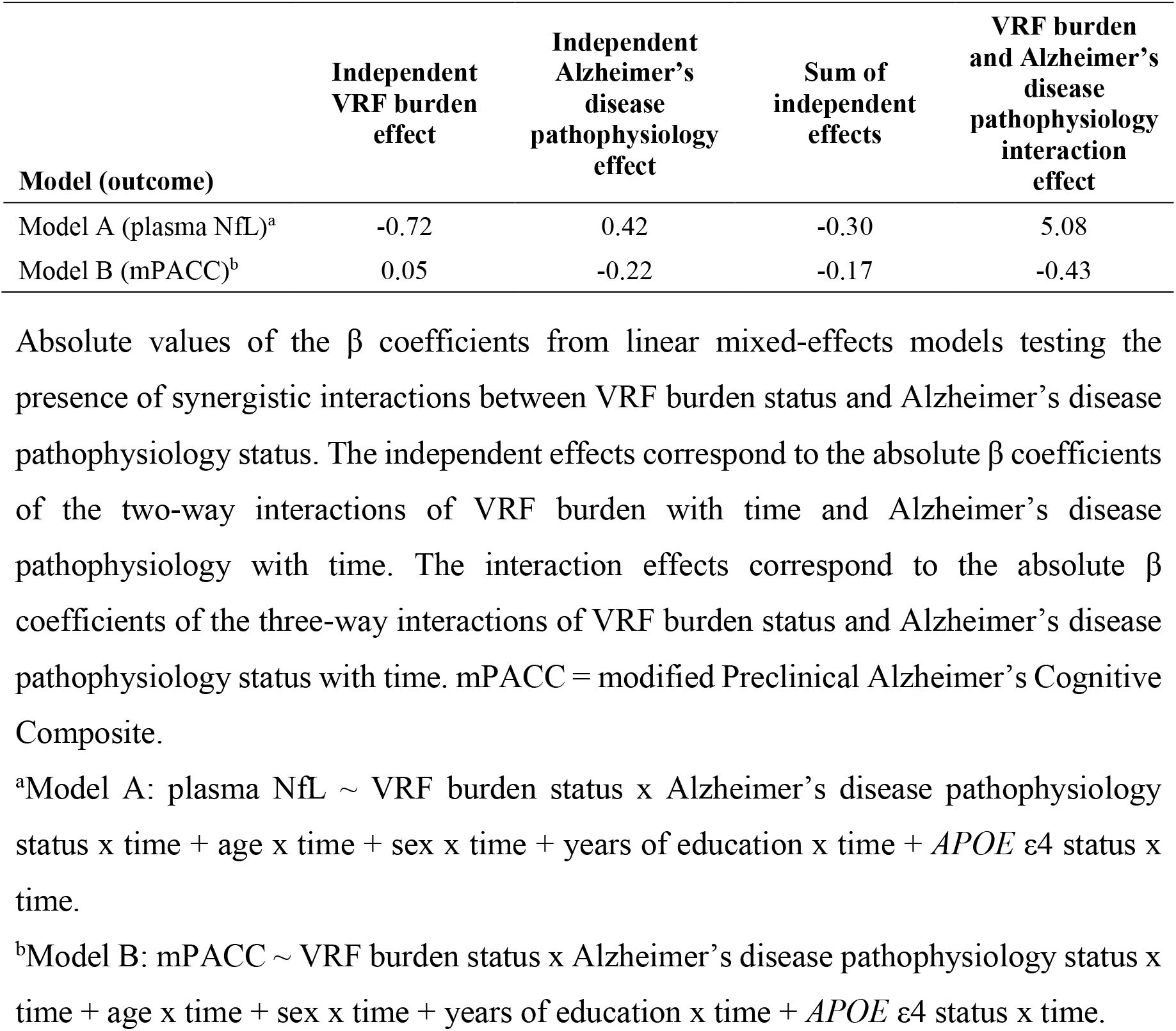
Independent and interactive effects of vascular risk factor (VRF) burden and Alzheimer’s disease pathophysiology on longitudinal plasma neurofilament light (NfL) and cognitive trajectories.

| **Model (outcome)** | **Independent VRF burden effect** | **Independent Alzheimer's disease pathophysiology effect** | **Sum of independent effects** | **VRF burden and Alzheimer's disease pathophysiology interaction effect** |
| --- | --- | --- | --- | --- |
| Model A (plasma NfL)<sup>a</sup> | -0.72 | 0.42 | -0.30 | 5.08 |
| Model B (mPACC)<sup>b</sup> | 0.05 | -0.22 | -0.17 | -0.43 |

Absolute values of the $\beta$ coefficients from linear mixed-effects models testing the presence of synergistic interactions between VRF burden status and Alzheimer's disease pathophysiology status. The independent effects correspond to the absolute $\beta$ coefficients of the two-way interactions of VRF burden with time and Alzheimer's disease pathophysiology with time. The interaction effects correspond to the absolute $\beta$ coefficients of the three-way interactions of VRF burden status and Alzheimer's disease pathophysiology status with time. mPACC = modified Preclinical Alzheimer's Cognitive Composite.
<sup>a</sup>Model A: plasma NfL ∼ VRF burden status x Alzheimer's disease pathophysiology status x time + age x time + sex x time + years of education x time + *APOE* ε4 status x time.
<sup>b</sup>Model B: mPACC ∼ VRF burden status x Alzheimer's disease pathophysiology status x time + age x time + sex x time + years of education x time + *APOE* ε4 status x time.

**Figure 1.**
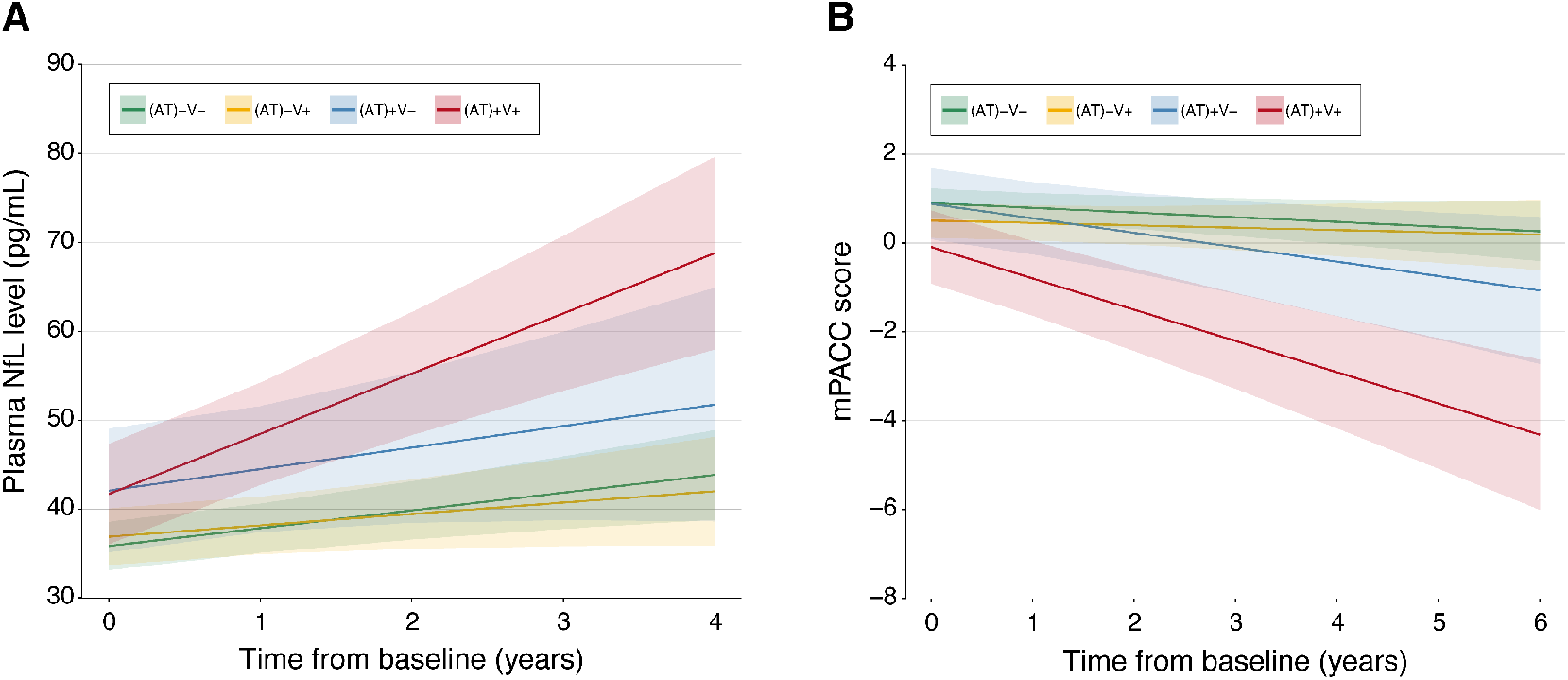
Elevated vascular risk factor (VRF) burden accelerates neurodegeneration and cognitive decline in subjects with preclinical Alzheimer’s disease. Mean predicted trajectories and 95% confidence intervals estimated from linear mixed-effects models according to baseline and VRF burden status and Alzheimer’s disease pathophysiology status. (**A**) Longitudinal neurodegeneration measured by plasma neurofilament light (NfL) in a 4-year follow-up period and (**B**) longitudinal cognitive trajectory indexed by the modified Preclinical Alzheimer’s Cognitive Composite (mPACC) in a 6-year follow-up period. Each model was adjusted for age, sex, years of education, apolipoprotein E ε4 status, and their interaction with time.

### 3.2. VRF burden and Alzheimer’s disease pathophysiology act synergistically on cognitive decline

Coefficients from LME models assessing the associations between VRF burden, Alzheimer’s disease pathophysiology, and longitudinal cognitive decline are shown in **Table 2**, Model B. Worse baseline cognitive performance was marginally associated with an elevated VRF burden (β = −0.39, *P* = 0.063). In contrast, no relation was detected with Alzheimer’s disease pathophysiology (*P* = 0.969) or with VRF burden x Alzheimer’s disease pathophysiology interaction (*P* = 0.312). Regarding cognitive trajectory, the three-way interaction (VRF burden x Alzheimer’s disease pathophysiology x time) was significant for predicting longitudinal cognitive decline (β = −0.43, *P* = 0.020), statistically supporting the notion that simultaneously having elevated VRF burden and preclinical Alzheimer’s disease accelerates the rates of cognitive decline more than the added impact of these conditions (i.e., synergy). Noteworthy, the interactive effect of VRF burden and Alzheimer’s disease pathophysiology was greater than the sum of their independent effects, confirming the presence of a synergistic relationship rather than the presence of additive effects (**Table 3**, Model B). Even though VRF burden was not associated with changes in cognition over time (i.e., VRF burden x time interaction term was not significant; *P* = 0.431), we detected a trend for the impact of preclinical Alzheimer’s disease on the rate of cognitive decline (i.e., Alzheimer’s disease pathophysiology x time interaction term was marginally significant; β = −0.22, *P* = 0.096). For results stratified by groups, see **Fig. 1B**.

### 3.3. VRF burden is not associated with Alzheimer’s disease CSF biomarkers

In LME models evaluating CSF Aβ_1-42_ and p-tau_181_ trajectories, neither the main effect of VRF burden nor the VRF burden x time interaction was significant (**Fig. 2A** and Model C in **Table 2** for CSF Aβ_1-42_ and **Fig. 2B** and Model D in **Table 2** for p-tau_181_; all *P* ≥ 0.314). Hence, VRF burden was not associated with baseline CSF Aβ_1-42_ and p-tau_181_ levels nor with changes in its levels over time.

**Figure 2.**
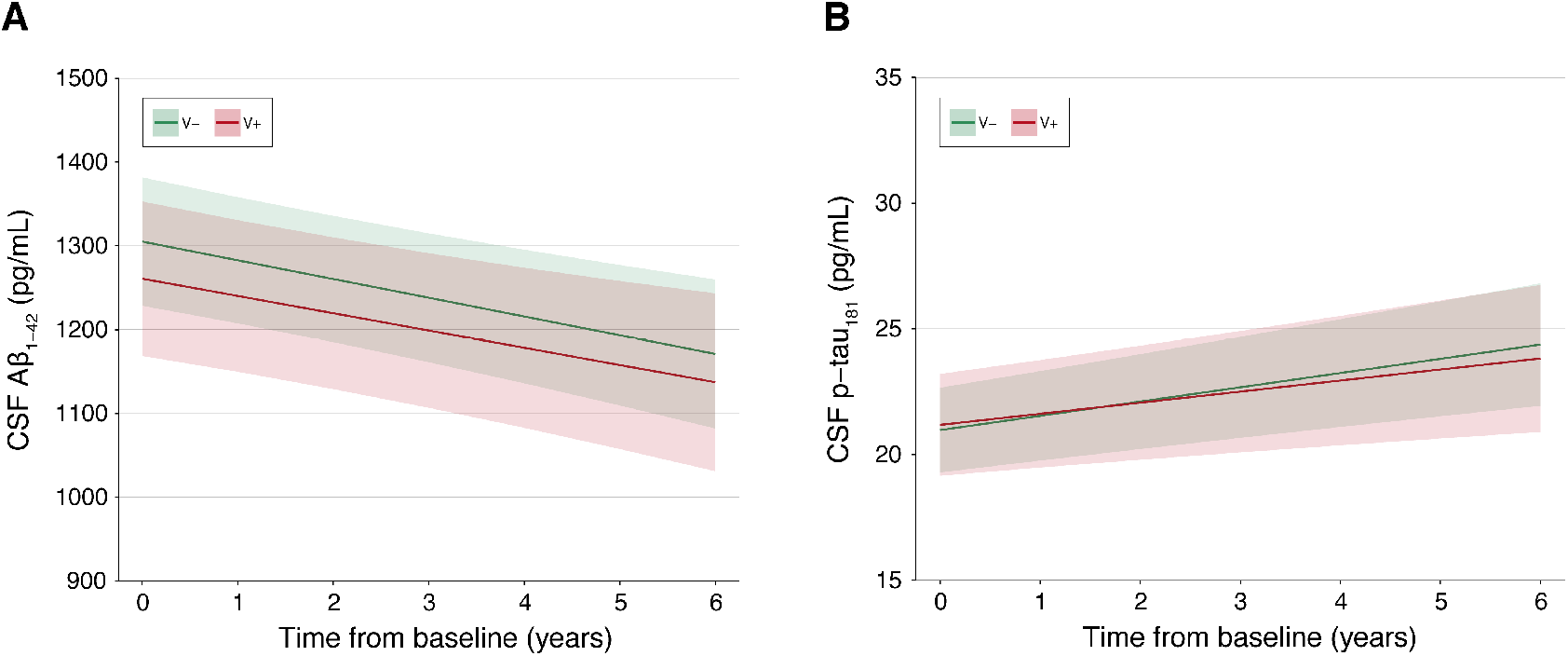
Vascular risk factor (VRF) burden is not associated with changes over time in cerebrospinal fluid (CSF) Aβ_1-42_ and p-tau_181_ levels. Mean predicted trajectories and 95% confidence intervals estimated from linear mixed-effects models according to baseline VRF burden status. (**A**) CSF Aβ_1-42_ longitudinal trajectory in a 6-year follow-up and (**B**) CSF p-tau_181_ longitudinal trajectory in a 6-year follow-up period. Each model was adjusted for age, sex, years of education, apolipoprotein E ε4 status, and their interaction with time. Aβ_1-42_ = amyloid-β_1-42_; p-tau_181_ = tau phosphorylated at threonine 181.

### 3.4. Increased risk of clinical progression in (AT)+V+ individuals

Kaplan-Meier curves demonstrate that the (AT)+V+ group presented a well-separated survival probability curve in a 6-year follow-up period, while the trajectories of the remaining groups [(AT)-V-, (AT)-V+, and (AT)+V-] were overlapping among themselves (**Fig. 3A**). The log-rank test showed a significant difference in estimated survival probabilities across the four groups (*P* < 0.0001). Cox-proportional hazards models corroborated that only the (AT)+V+ group had a significantly higher risk of clinical progression compared to the reference group [(AT)-V-] in 6 years (adjusted HR = 3.5, *P* < 0.001; **Table 4**). Post-hoc comparisons demonstrated that (AT)+V+ group presented a significantly higher risk of clinical progression in comparison to (AT)-V- (*P* = 0.005) and (AT)-V+ (*P* = 0.012) groups, whereas it did not reach statistical significance in comparison to the (AT)+V-group (*P* = 0.127). Additionally, no differences were observed in pairwise contrast analysis among the (AT)-V-, (AT)-V+, and (AT)+V-groups. In further analyses not adjusting for potential confounders, we observed similar findings (**Supplementary Table 4**). To better understand the unexpected slow conversion in the (AT)+V-group, we conducted a few exploratory analyses testing clinical deterioration detected by the CDR-SB scale in the whole sample (**Fig. 3B**) and in individuals under 75 years (**Fig. 3C**), as well as survival probabilities using WMH status (instead of VRF burden status) for clinical conversion (**Fig. 3D**) and for changes in the CDR-SB scale (**Fig. 3E**) in the whole sample. The reduced number of conversions is a limiting factor that does not allow definitive conclusions. This issue is properly discussed in the limitation section and deserves further investigation.

**Table 4.** Adjusted Hazard Ratios of clinical progression according to vascular risk factor (VRF) burden and Alzheimer’s disease pathophysiology status.

|  | **Adjusted Hazard Ratio (95% CI)** | **P-value** |
| --- | --- | --- |
| (AT)-V- | =1 (reference) | - |
| (AT)-V+ | 1.2 (0.7 to 2.1) | 0.555 |
| (AT)+V- | 0.97 (0.3 to 2.9) | 0.963 |
| (AT)+V+ | 3.5 (1.7 to 7.3) | < 0.001 |

Cox proportional hazards models evaluating risk of conversion to cognitive impairment (i.e. mild cognitive impairment or dementia) in 6 years according to VRF burden status and Alzheimer's disease pathophysiology status. Model was fitted with potential confounders (age, years of education, sex, and apolipoprotein E ε4 status). For this analysis, the (AT)-V- group was considered as the reference group. Post-hoc comparisons regarding the risk of clinical progression across the four groups are described in the text. CI = confidence interval.

**Figure 3.**
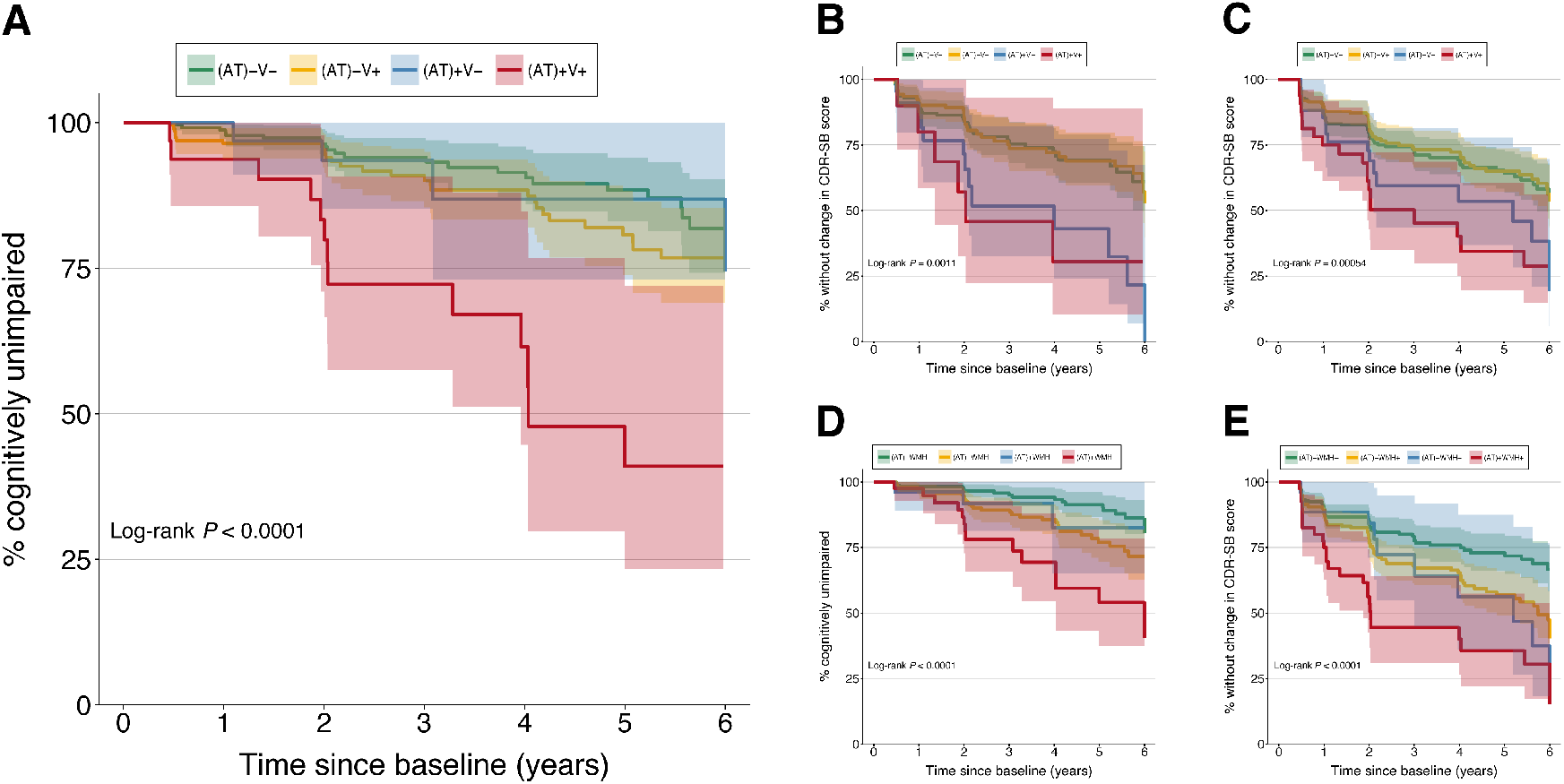
Individuals with preclinical Alzheimer’s disease and elevated vascular risk factor (VRF) burden have a greater risk of clinical progression. (**A**) Conversion to cognitive impairment [i.e., mild cognitive impairment (MCI) or dementia] in the whole sample according to baseline VRF burden status and Alzheimer’s disease pathophysiology status. (**B**) Changes in the clinical dementia rating scale sum of boxes (CDR-SB) scale in the whole sample according to baseline VRF burden status and Alzheimer’s disease pathophysiology status. (**C**) Changes in the CDR-SB scale in individuals under 75 years according to baseline VRF burden status and Alzheimer’s disease pathophysiology status. (**D**) Conversion to cognitive impairment (i.e., MCI or dementia) in the whole sample according to baseline white matter hyperintensities (WMH) status and Alzheimer’s disease pathophysiology status. (**E**) Changes in the CDR-SB scale in the whole sample according to baseline WMH status and Alzheimer’s disease pathophysiology status. Kaplan-Meier curves with 95% confidence intervals display survival probability for each group. Significance level obtained from the log-rank test is also reported.

## 4. Discussion

In the present study, we showed that VRF burden interacts synergistically with Alzheimer’s disease pathophysiology to drive longitudinal increases in plasma NfL levels, as well as longitudinal decline in mPACC scores in CU individuals. Nevertheless, VRF burden alone was not associated with changes in CSF Alzheimer’s disease biomarkers (CSF Aβ_1-42_ and p-tau_181_). Additionally, we demonstrated that the presence of both elevated VRF burden and preclinical Alzheimer’s disease in CU individuals is associated with higher progression rates to MCI or dementia. These findings suggest that the impact of VRF burden on neurodegeneration and cognitive decline varies according to Alzheimer’s disease pathophysiology status in CU subjects.

Here, we observed that VRF burden and Alzheimer’s disease pathophysiology were synergistically associated with longitudinal neurodegeneration measured by plasma NfL. Even though it focused on another neurodegeneration biomarker, a previous study reported that CU individuals with an elevated Framingham Risk Score presented higher rates of increase in CSF t-tau levels when abnormal to Aβ and tau biomarkers at baseline.^43^ Given that the association was restricted to the A+T+ group, this finding further supports the notion of an interactive association between vascular risk and Alzheimer’s disease pathophysiology. Also, our results are in agreement with cross-sectional neuroimaging evidence that vascular risk and Aβ burden were interactively associated with lower cortical thickness in Alzheimer’s disease vulnerable (posterior) brain regions in CU and mildly impaired subjects.^44^ Taken together, this complimentary evidence highlights the importance of considering the VRF burden when studying the processes underlying brain damage in preclinical Alzheimer’s disease.

The notion that VRF burden and Alzheimer’s disease pathophysiology interact to promote cognitive impairment is also supported by recently published evidence in the literature. In participants from the Harvard Aging Brain Study (HABS), it was observed a synergistic association of vascular risk and Aβ burden with prospective cognitive decline in CU elderlies.^22^ Similarly, another longitudinal study including CU older adults demonstrated that an increased Framingham Risk Score was associated with higher rates of cognitive decline only in the A+T+ group, indicating an interactive effect.^43^ Notwithstanding, divergent results have also been reported. A recent investigation evaluating CU participants from the Biomarkers for Older Controls at Risk for Dementia (BIOCARD) study concluded that midlife vascular risk and Alzheimer’s disease pathophysiology – defined by CSF biomarkers – presented additive rather than synergistic effects on cognitive decline over a mean follow-up of 13.9 years.^20^ Although assessing VRFs in midlife and having a longer follow-up is a clear strength of this work, other factors could account for the divergent results, such as vascular risk assessment (dichotomization by 0 or ≥ 1 evaluating the following conditions: hypertension, hypercholesterolemia, diabetes, smoking, and obesity) and cutpoints used for Alzheimer’s disease biomarkers (based on tertiles calculated considering midlife Alzheimer’s disease biomarker concentrations). Together, these findings reinforce that, even without a clear consensus in the literature, VRFs and Alzheimer’s disease pathophysiology often coexist and play pivotal roles in brain aging.

Furthermore, our results suggest that VRF burden does not directly impact Alzheimer’s disease pathophysiology. Previous studies have reported inconsistent results concerning the links between VRFs and Alzheimer’s disease pathophysiology. While some investigations found that VRFs were cross-sectionally associated either with higher Aβ or tau burden,^18, 19^ most evidence is against the presence of such relations.^12, 22, 23^ Nonetheless, contrary to our results, a recent study showed that the cumulative number of midlife VRFs was associated with late-life Aβ deposition,^16^ as well as that high vascular risk and elevated Aβ burden at baseline were interactively associated with increased tau PET signal in the inferior temporal cortex measured nearly three years later.^17^ On the other hand, our finding that the VRF burden does not present short-term influence on Alzheimer’s disease pathophysiology is in line with recent studies evaluating the associations between composite vascular risk scores – calculated based on the cumulative number of vascular risk factors or the Framingham Risk Score – and longitudinal changes in the CSF Aβ_1-42_ and p-tau_181_ levels.^20, 21, 43^ To rule out or confirm a possible association between VRF burden and Alzheimer’s disease pathophysiology, further cohort studies with longer follow-up and increased sample size, ideally using neuroimaging biomarkers, are needed. Lastly, it is vital to acknowledge the possibility of differential mechanisms and weights specific for each VRF, which was beyond the scope of the present investigation.

Not rarely do individuals without cognitive impairment present Alzheimer’s disease pathophysiology,^45–49^ highlighting the role of both resilience mechanisms and concomitant pathological processes in the clinical expression of Alzheimer’s disease. In this context, it has been proposed that vascular dysfunction has an early role in Alzheimer’s disease progression.^50^ There are different potential mechanisms by which vascular factors contribute to cognitive impairment and dementia, such as reduction in cerebral blood flow and hypoxia, blood-brain barrier (BBB) breakdown, endothelial dysfunction, systemic inflammation and oxidative stress, and disruption of trophic coupling.^51^ Our results support the notion that CU individuals exposed to higher VRFs might have a decreased threshold for cognitive decline and neurodegeneration induced by Alzheimer’s disease pathophysiology. As previously suggested, a possible explanation is that VRFs influence the progression of Alzheimer’s disease through the promotion of cerebrovascular injuries rather than through a direct effect on Alzheimer’s disease pathophysiology.^23^ Additionally, BBB breakdown, an important feature in early Alzheimer’s disease^52–55^ and a potential biomarker of cognitive dysfunction,^56^ is associated with both Alzheimer’s disease pathophysiology and VRFs, but at different molecular levels, stressing the role of brain vasculature in cognitive impairment. Hence, VRFs appear to impact brain resilience mechanisms against the deposition of Aβ-containing extracellular neuritic plaques and tau-containing neurofibrillary tangles.

At the moment, there is no approved pharmacological treatment that can unquestionably stop or delay Alzheimer’s disease clinical deterioration. Since Alzheimer’s disease is a multifactorial disease, it is reasonable to consider that an effective therapy would need to have multiple targets, not only Aβ and tau accumulation. Also, given that Alzheimer’s disease pathophysiology starts to accumulate many years before the onset of clinical symptoms,^57^ new clinical trials often focus on asymptomatic individuals presenting biomarker evidence of Aβ and tau pathologies (i.e., preclinical stages of Alzheimer’s disease).^1, 2^ The findings from the present work corroborate that the development of therapies targeting both Alzheimer’s disease pathophysiology and VRFs in Alzheimer’s disease preclinical stages could potentiate treatment response.

Similar to ADNI’s enrollment criteria, clinical trials for Alzheimer’s disease usually do not include individuals with an elevated cerebrovascular burden. To this end, enrollment criteria commonly exclude subjects with Hachinski ischemic scores equal or greater than four and having brain MRI lesions.^58–60^ Excluding patients with significant cerebrovascular disease could be seen as a limitation of the present study as it makes the cohort underrepresented in terms of vascular pathology. However, we could still observe an interactive association of VRF burden with Alzheimer’s disease pathophysiology on longitudinal neurodegeneration and cognitive decline in CU individuals. These results suggest that future clinical trials in individuals with preclinical Alzheimer’s disease should not exclude a possible vascular contribution to Alzheimer’s disease progression by merely using the aforementioned enrollment criteria. On the other hand, a potential approach for studies aiming to solely focus on Alzheimer’s disease pathophysiology is to further apply the composite VRF burden score used in the present work.

Even though cognition is the primary outcome of interest in disease-modifying drug trials, individuals with preclinical Alzheimer’s disease can stay cognitively stable over many years.^4^ Therefore, an important limitation for the performance of these trials is the need for high sample sizes and extended follow-ups. In this context, the use of surrogate markers of disease progression can be a useful alternative. Besides being noninvasive and accessible, plasma NfL has been shown to be a robust neurodegeneration marker to monitor Alzheimer’s disease progression.^31, 61^ Nonetheless, to the best of our knowledge, no previous study has longitudinally appraised the associations of VRF burden and Alzheimer’s disease pathophysiology with plasma NfL trajectories. Similar to the pattern observed for cognition, our results showed that the synergy between VRF burden and Alzheimer’s disease pathophysiology led to longitudinal increases in plasma NfL. Together, these findings suggest that plasma NfL may be used as a surrogate to track therapeutic response in trials targeting vascular risk factors and Alzheimer’s disease pathophysiology.

From the perspective of preventive strategies, a recent report estimated that modifiable risk factors – including VRFs such as hypertension, smoking, and diabetes – are associated with nearly 40% of dementia cases, which could be potentially prevented or delayed.^62^ In fact, a recent study has suggested that the reduction in dementia incidence in participants of the Framingham Heart Study over the past three decades could be, at least in part, a consequence of improved management of VRFs.^63^ Additionally, the multicenter Finnish Geriatric Intervention Study to Prevent Cognitive Impairment and Disability (FINGER) trial has shown that a multidomain lifestyle intervention – which involved controlling VRFs – provided benefits for cognitive performance in a population of older subjects at increased risk for cognitive decline and dementia.^64^ In accordance with the literature evidence, the data presented in this investigation reinforces the importance of developing preventive strategies for reducing dementia societal burden, which must include maintaining a good vascular health.

Some limitations need to be highlighted to appropriately interpret our results. First, the ADNI study involves a selective population of highly educated and mostly white volunteers, which might further jeopardize the generalizability of our results, especially because different ethnic groups are not homogenously affected by VRFs.^65–67^ Second, it is also necessary to consider the limited sample size, mainly of participants with preclinical Alzheimer’s disease. This issue is probably the reason for the wide confidence intervals in time-to-event analysis. Third, we did not consider the age of onset of the VRFs. Some VRFs may have an age-dependent effect – such as midlife and late-life exposures being differentially related to the risk of dementia and specifically Alzheimer’s disease.^11, 68–71^ Thus, we cannot determine whether our findings would differ considering only exposures that started in midlife or late-life. Fourth, the presence of VRFs was determined based on previous diagnosis and use of medications information collected in clinical interviews rather than diagnosis performed at study entry with objective measurements, potentially being a source of bias. Fifth, previous studies demonstrated that CSF Aβ_1-42/1-40_ ratio better predict abnormal brain Aβ accumulation in comparison to CSF Aβ_1-42_.^72^ However, it was not possible to determine Aβ positivity using the CSF Aβ_1-42/1-40_ ratio in the present study due to limited CSF Aβ_1-40_ data available.

In conclusion, we demonstrated that VRF burden and biomarker evidence of Alzheimer’s disease pathophysiology are synergistically associated with neurodegeneration and cognitive deterioration in CU individuals, whereas VRF burden does not influence Aβ and tau pathologies. Our results provide additional evidence for the performance of clinical trials targeting VRFs and Alzheimer’s disease pathophysiology. Additionally, these trials could have advantages from using plasma NfL as a surrogate to track therapeutic response. Importantly, further community-based studies with longer follow-up and bigger sample sizes are needed to confirm our findings.

## Supporting information

STROBE statement

## Data Availability

Data used in the preparation of the present manuscript is publicly available for download at http://adni.loni.usc.edu. If needed for replication purposes, derived data used in our analysis will be made available upon reasonable request.

## Abbreviations

ADAS-Cog: Alzheimer’s Disease Assessment Scale – Cognitive Subscale
ADNI: Alzheimer’s Disease Neuroimaging Initiative
*APOE* ε4: Apolipoprotein E ε4
Aβ_1-42_: amyloid-β_1-42_
BBB: blood-brain barrier
BIOCARD: Biomarkers for Older Controls at Risk for Dementia
(CDR-SB): clinical dementia rating scale sum of boxes
CU: cognitively unimpaired
FINGER: Finnish Geriatric Intervention Study to Prevent Cognitive Impairment and Disability
HABS: Harvard Aging Brain Study
HR: Hazard Ratio
LME: linear mixed-effects
MCI: mild cognitive impairment
MMSE: Mini-Mental State Examination
mPACC: modified version of Preclinical Alzheimer’s Cognitive Composite
NfL: neurofilament light
NIA-AA: National Institute on Aging and the Alzheimer’s Association
p-tau_181_: tau phosphorylated at threonine 181
SD: standard deviation
Simoa: single molecule array
t-tau: total tau
TIA: transient ischemic attack
VRF: vascular risk factor
VIF: variance inflation factor
WMH: white matter hyperintensities.

## Acknowledgements

Data collection and sharing for this project was funded by the Alzheimer’s Disease Neuroimaging Initiative (ADNI) (National Institutes of Health Grant U01 AG024904) and DOD ADNI (Department of Defense award number W81XWH-12-2-0012). ADNI is funded by the National Institute on Aging, the National Institute of Biomedical Imaging and Bioengineering, and through generous contributions from the following: AbbVie, Alzheimer’s Association; Alzheimer’s Drug Discovery Foundation; Araclon Biotech; BioClinica, Inc.; Biogen; Bristol-Myers Squibb Company; CereSpir, Inc.; Cogstate; Eisai Inc.; Elan Pharmaceuticals, Inc.; Eli Lilly and Company; EuroImmun; F. Hoffmann-La Roche Ltd and its affiliated company Genentech, Inc.; Fujirebio; GE Healthcare; IXICO Ltd.; Janssen Alzheimer Immunotherapy Research & Development, LLC.; Johnson & Johnson Pharmaceutical Research & Development LLC.; Lumosity; Lundbeck; Merck & Co., Inc.; Meso Scale Diagnostics, LLC.; NeuroRx Research; Neurotrack Technologies; Novartis Pharmaceuticals Corporation; Pfizer Inc.; Piramal Imaging; Servier; Takeda Pharmaceutical Company; and Transition Therapeutics. The Canadian Institutes of Health Research is providing funds to support ADNI clinical sites in Canada. Private sector contributions are facilitated by the Foundation for the National Institutes of Health (www.fnih.org). The grantee organization is the Northern California Institute for Research and Education, and the study is coordinated by the Alzheimer’s Therapeutic Research Institute at the University of Southern California. ADNI data are disseminated by the Laboratory for Neuro Imaging at the University of Southern California.

## References

1. Sperling RA, Rentz DM, Johnson KA, et al. The A4 study: stopping AD before symptoms begin? Sci Transl Med. Mar 19 2014;6(228):228fs13. doi:10.1126/scitranslmed.3007941

2. Cummings J, Lee G, Zhong K, Fonseca J, Taghva K. Alzheimer’s disease drug development pipeline: 2021. Alzheimers Dement (N Y). 2021;7(1):e12179. doi:10.1002/trc2.12179

3. Jack CR, Jr., Bennett DA, Blennow K, et al. NIA-AA Research Framework: Toward a biological definition of Alzheimer’s disease. Alzheimers Dement. Apr 2018;14(4):535–562. doi:10.1016/j.jalz.2018.02.018

4. Dubois B, Villain N, Frisoni GB, et al. Clinical diagnosis of Alzheimer’s disease: recommendations of the International Working Group. Lancet Neurol. Jun 2021;20(6):484–496. doi:10.1016/S1474-4422(21)00066-1

5. Masters CL, Bateman R, Blennow K, Rowe CC, Sperling RA, Cummings JL. Alzheimer’s disease. Nat Rev Dis Primers. Oct 15 2015;1:15056. doi:10.1038/nrdp.2015.56

6. Luchsinger JA, Reitz C, Honig LS, Tang MX, Shea S, Mayeux R. Aggregation of vascular risk factors and risk of incident Alzheimer disease. Neurology. Aug 23 2005;65(4):545–51. doi:10.1212/01.wnl.0000172914.08967.dc

7. Kivipelto M, Helkala EL, Laakso MP, et al. Midlife vascular risk factors and Alzheimer’s disease in later life: longitudinal, population based study. BMJ. Jun 16 2001;322(7300):1447–51. doi:10.1136/bmj.322.7300.1447

8. Ott A, Stolk RP, van Harskamp F, Pols HA, Hofman A, Breteler MM. Diabetes mellitus and the risk of dementia: The Rotterdam Study. Neurology. Dec 10 1999;53(9):1937–42. doi:10.1212/wnl.53.9.1937

9. Profenno LA, Porsteinsson AP, Faraone SV. Meta-analysis of Alzheimer’s disease risk with obesity, diabetes, and related disorders. Biol Psychiatry. Mar 15 2010;67(6):505–12. doi:10.1016/j.biopsych.2009.02.013

10. Anstey KJ, von Sanden C, Salim A, O’Kearney R. Smoking as a risk factor for dementia and cognitive decline: a meta-analysis of prospective studies. Am J Epidemiol. Aug 15 2007;166(4):367–78. doi:10.1093/aje/kwm116

11. Anstey KJ, Ashby-Mitchell K, Peters R. Updating the Evidence on the Association between Serum Cholesterol and Risk of Late-Life Dementia: Review and Meta-Analysis. J Alzheimers Dis. 2017;56(1):215–228. doi:10.3233/JAD-160826

12. Bangen KJ, Nation DA, Delano-Wood L, et al. Aggregate effects of vascular risk factors on cerebrovascular changes in autopsy-confirmed Alzheimer’s disease. Alzheimers Dement. Apr 2015;11(4):394–403 e1. doi:10.1016/j.jalz.2013.12.025

13. Nation DA, Delano-Wood L, Bangen KJ, et al. Antemortem pulse pressure elevation predicts cerebrovascular disease in autopsy-confirmed Alzheimer’s disease. J Alzheimers Dis. 2012;30(3):595–603. doi:10.3233/JAD-2012-111697

14. Snowdon DA, Greiner LH, Mortimer JA, Riley KP, Greiner PA, Markesbery WR. Brain infarction and the clinical expression of Alzheimer disease. The Nun Study. JAMA. Mar 12 1997;277(10):813–7.

15. Attems J, Jellinger KA. The overlap between vascular disease and Alzheimer’s disease--lessons from pathology. BMC Med. Nov 11 2014;12:206. doi:10.1186/s12916-014-0206-2

16. Gottesman RF, Schneider AL, Zhou Y, et al. Association Between Midlife Vascular Risk Factors and Estimated Brain Amyloid Deposition. JAMA. Apr 11 2017;317(14):1443–1450. doi:10.1001/jama.2017.3090

17. Rabin JS, Yang HS, Schultz AP, et al. Vascular Risk and beta-Amyloid Are Synergistically Associated with Cortical Tau. Ann Neurol. Feb 2019;85(2):272–279. doi:10.1002/ana.25399

18. Vemuri P, Lesnick TG, Przybelski SA, et al. Age, vascular health, and Alzheimer disease biomarkers in an elderly sample. Ann Neurol. Nov 2017;82(5):706–718. doi:10.1002/ana.25071

19. Kobe T, Gonneaud J, Pichet Binette A, et al. Association of Vascular Risk Factors With beta-Amyloid Peptide and Tau Burdens in Cognitively Unimpaired Individuals and Its Interaction With Vascular Medication Use. JAMA Netw Open. Feb 5 2020;3(2):e1920780. doi:10.1001/jamanetworkopen.2019.20780

20. Pettigrew C, Soldan A, Wang J, et al. Association of midlife vascular risk and AD biomarkers with subsequent cognitive decline. Neurology. Dec 8 2020;95(23):e3093–e3103. doi:10.1212/WNL.0000000000010946

21. Lo RY, Jagust WJ, Alzheimer’s Disease Neuroimaging I. Vascular burden and Alzheimer disease pathologic progression. Neurology. Sep 25 2012;79(13):1349–55. doi:10.1212/WNL.0b013e31826c1b9d

22. Rabin JS, Schultz AP, Hedden T, et al. Interactive Associations of Vascular Risk and beta-Amyloid Burden With Cognitive Decline in Clinically Normal Elderly Individuals: Findings From the Harvard Aging Brain Study. JAMA Neurol. Sep 1 2018;75(9):1124–1131. doi:10.1001/jamaneurol.2018.1123

23. Chui HC, Zheng L, Reed BR, Vinters HV, Mack WJ. Vascular risk factors and Alzheimer’s disease: are these risk factors for plaques and tangles or for concomitant vascular pathology that increases the likelihood of dementia? An evidence-based review. Alzheimers Res Ther. Jan 4 2012;4(1):1. doi:10.1186/alzrt98

24. Petersen RC, Aisen PS, Beckett LA, et al. Alzheimer’s Disease Neuroimaging Initiative (ADNI): clinical characterization. Neurology. Jan 19 2010;74(3):201–9. doi:10.1212/WNL.0b013e3181cb3e25

25. Lloyd-Jones DM, Leip EP, Larson MG, et al. Prediction of lifetime risk for cardiovascular disease by risk factor burden at 50 years of age. Circulation. Feb 14 2006;113(6):791–8. doi:10.1161/CIRCULATIONAHA.105.548206

26. Berry JD, Dyer A, Cai X, et al. Lifetime risks of cardiovascular disease. N Engl J Med. Jan 26 2012;366(4):321–9. doi:10.1056/NEJMoa1012848

27. Bittner T, Zetterberg H, Teunissen CE, et al. Technical performance of a novel, fully automated electrochemiluminescence immunoassay for the quantitation of beta-amyloid (1-42) in human cerebrospinal fluid. Alzheimers Dement. May 2016;12(5):517–26. doi:10.1016/j.jalz.2015.09.009

28. Lifke V, Kollmorgen G, Manuilova E, et al. Elecsys((R)) Total-Tau and Phospho-Tau (181P) CSF assays: Analytical performance of the novel, fully automated immunoassays for quantification of tau proteins in human cerebrospinal fluid. Clin Biochem. Oct 2019;72:30–38. doi:10.1016/j.clinbiochem.2019.05.005

29. Ingala S, De Boer C, Masselink LA, et al. Application of the ATN classification scheme in a population without dementia: Findings from the EPAD cohort. Alzheimers Dement. Jul 2021;17(7):1189–1204. doi:10.1002/alz.12292

30. Mattsson N, Andreasson U, Zetterberg H, Blennow K, Alzheimer’s Disease Neuroimaging I. Association of Plasma Neurofilament Light With Neurodegeneration in Patients With Alzheimer Disease. JAMA Neurol. May 1 2017;74(5):557–566. doi:10.1001/jamaneurol.2016.6117

31. Mattsson N, Cullen NC, Andreasson U, Zetterberg H, Blennow K. Association Between Longitudinal Plasma Neurofilament Light and Neurodegeneration in Patients With Alzheimer Disease. JAMA Neurol. Jul 1 2019;76(7):791–799. doi:10.1001/jamaneurol.2019.0765

32. Schwarz C, Fletcher E, DeCarli C, Carmichael O. Fully-automated white matter hyperintensity detection with anatomical prior knowledge and without FLAIR. Inf Process Med Imaging. 2009;21:239–51. doi:10.1007/978-3-642-02498-6_20

33. DeCarli C, Maillard P, Fletcher E. Four Tissue Segmentation in ADNI II. Accessed 27 October 2021, https://files.alz.washington.edu/documentation/adni-proto.pdf

34. Donohue MC, Sperling RA, Salmon DP, et al. The preclinical Alzheimer cognitive composite: measuring amyloid-related decline. JAMA Neurol. Aug 2014;71(8):961–70. doi:10.1001/jamaneurol.2014.803

35. Mattsson-Carlgren N, Janelidze S, Palmqvist S, et al. Longitudinal plasma p-tau217 is increased in early stages of Alzheimer’s disease. Brain. Dec 5 2020;143(11):3234–3241. doi:10.1093/brain/awaa286

36. Hansson O, Seibyl J, Stomrud E, et al. CSF biomarkers of Alzheimer’s disease concord with amyloid-beta PET and predict clinical progression: A study of fully automated immunoassays in BioFINDER and ADNI cohorts. Alzheimers Dement. Nov 2018;14(11):1470–1481. doi:10.1016/j.jalz.2018.01.010

37. Blennow K, Shaw LM, Stomrud E, et al. Predicting clinical decline and conversion to Alzheimer’s disease or dementia using novel Elecsys Abeta(1-42), pTau and tTau CSF immunoassays. Sci Rep. Dec 13 2019;9(1):19024. doi:10.1038/s41598-019-54204-z

38. Berrington de González A, Cox DR. Interpretation of interaction: A review. The Annals of Applied Statistics. 2007;1(2):371–385. doi:10.1214/07-aoas124

39. Slinker BK. The statistics of synergism. J Mol Cell Cardiol. Apr 1998;30(4):723–31. doi:10.1006/jmcc.1998.0655

40. Therriault J, Benedet AL, Pascoal TA, et al. APOEepsilon4 potentiates the relationship between amyloid-beta and tau pathologies. Mol Psychiatry. Mar 11 2020;doi:10.1038/s41380-020-0688-6

41. Wagner M, Helmer C, Tzourio C, Berr C, Proust-Lima C, Samieri C. Evaluation of the Concurrent Trajectories of Cardiometabolic Risk Factors in the 14 Years Before Dementia. JAMA Psychiatry. Oct 1 2018;75(10):1033–1042. doi:10.1001/jamapsychiatry.2018.2004

42. Steyerberg EW. Clinical prediction models : a practical approach to development, validation, and updating. Springer; 2019.

43. Bos I, Vos SJB, Schindler SE, et al. Vascular risk factors are associated with longitudinal changes in cerebrospinal fluid tau markers and cognition in preclinical Alzheimer’s disease. Alzheimers Dement. Sep 2019;15(9):1149–1159. doi:10.1016/j.jalz.2019.04.015

44. Villeneuve S, Reed BR, Madison CM, et al. Vascular risk and Abeta interact to reduce cortical thickness in AD vulnerable brain regions. Neurology. Jul 1 2014;83(1):40–7. doi:10.1212/WNL.0000000000000550

45. Bennett DA, Schneider JA, Arvanitakis Z, et al. Neuropathology of older persons without cognitive impairment from two community-based studies. Neurology. Jun 27 2006;66(12):1837–44. doi:10.1212/01.wnl.0000219668.47116.e6

46. Price JL, Morris JC. Tangles and plaques in nondemented aging and “preclinical” Alzheimer’s disease. Ann Neurol. Mar 1999;45(3):358–68. doi:10.1002/1531-8249(199903)45:3<358::aid-ana12>3.0.co;2-x

47. Iacono D, Resnick SM, O’Brien R, et al. Mild cognitive impairment and asymptomatic Alzheimer disease subjects: equivalent beta-amyloid and tau loads with divergent cognitive outcomes. J Neuropathol Exp Neurol. Apr 2014;73(4):295–304. doi:10.1097/NEN.0000000000000052

48. Perez-Nievas BG, Stein TD, Tai HC, et al. Dissecting phenotypic traits linked to human resilience to Alzheimer’s pathology. Brain. Aug 2013;136(Pt 8):2510–26. doi:10.1093/brain/awt171

49. Lowe VJ, Bruinsma TJ, Min HK, et al. Elevated medial temporal lobe and pervasive brain tau-PET signal in normal participants. Alzheimers Dement (Amst). 2018;10:210–216. doi:10.1016/j.dadm.2018.01.005

50. Iturria-Medina Y, Sotero RC, Toussaint PJ, Mateos-Perez JM, Evans AC, Alzheimer’s Disease Neuroimaging I. Early role of vascular dysregulation on late-onset Alzheimer’s disease based on multifactorial data-driven analysis. Nat Commun. Jun 21 2016;7:11934. doi:10.1038/ncomms11934

51. Zlokovic BV, Gottesman RF, Bernstein KE, et al. Vascular contributions to cognitive impairment and dementia (VCID): A report from the 2018 National Heart, Lung, and Blood Institute and National Institute of Neurological Disorders and Stroke Workshop. Alzheimers Dement. Dec 2020;16(12):1714–1733. doi:10.1002/alz.12157

52. van de Haar HJ, Burgmans S, Jansen JF, et al. Blood-Brain Barrier Leakage in Patients with Early Alzheimer Disease. Radiology. Nov 2016;281(2):527–535. doi:10.1148/radiol.2016152244

53. van de Haar HJ, Jansen JFA, van Osch MJP, et al. Neurovascular unit impairment in early Alzheimer’s disease measured with magnetic resonance imaging. Neurobiol Aging. Sep 2016;45:190–196. doi:10.1016/j.neurobiolaging.2016.06.006

54. Montagne A, Barnes SR, Sweeney MD, et al. Blood-brain barrier breakdown in the aging human hippocampus. Neuron. Jan 21 2015;85(2):296–302. doi:10.1016/j.neuron.2014.12.032

55. Sweeney MD, Sagare AP, Zlokovic BV. Blood-brain barrier breakdown in Alzheimer disease and other neurodegenerative disorders. Nat Rev Neurol. Mar 2018;14(3):133–150. doi:10.1038/nrneurol.2017.188

56. Nation DA, Sweeney MD, Montagne A, et al. Blood-brain barrier breakdown is an early biomarker of human cognitive dysfunction. Nat Med. Feb 2019;25(2):270–276. doi:10.1038/s41591-018-0297-y

57. Sperling RA, Aisen PS, Beckett LA, et al. Toward defining the preclinical stages of Alzheimer’s disease: recommendations from the National Institute on Aging-Alzheimer’s Association workgroups on diagnostic guidelines for Alzheimer’s disease. Alzheimers Dement. May 2011;7(3):280–92. doi:10.1016/j.jalz.2011.03.003

58. Honig LS, Vellas B, Woodward M, et al. Trial of Solanezumab for Mild Dementia Due to Alzheimer’s Disease. N Engl J Med. Jan 25 2018;378(4):321–330. doi:10.1056/NEJMoa1705971

59. Mintun MA, Lo AC, Duggan Evans C, et al. Donanemab in Early Alzheimer’s Disease. N Engl J Med. May 6 2021;384(18):1691–1704. doi:10.1056/NEJMoa2100708

60. Ostrowitzki S, Lasser RA, Dorflinger E, et al. A phase III randomized trial of gantenerumab in prodromal Alzheimer’s disease. Alzheimers Res Ther. Dec 8 2017;9(1):95. doi:10.1186/s13195-017-0318-y

61. Benedet AL, Leuzy A, Pascoal TA, et al. Stage-specific links between plasma neurofilament light and imaging biomarkers of Alzheimer’s disease. Brain. Dec 1 2020;143(12):3793–3804. doi:10.1093/brain/awaa342

62. Livingston G, Huntley J, Sommerlad A, et al. Dementia prevention, intervention, and care: 2020 report of the Lancet Commission. Lancet. Aug 8 2020;396(10248):413–446. doi:10.1016/S0140-6736(20)30367-6

63. Satizabal CL, Beiser AS, Chouraki V, Chene G, Dufouil C, Seshadri S. Incidence of Dementia over Three Decades in the Framingham Heart Study. N Engl J Med. Feb 11 2016;374(6):523–32. doi:10.1056/NEJMoa1504327

64. Rosenberg A, Ngandu T, Rusanen M, et al. Multidomain lifestyle intervention benefits a large elderly population at risk for cognitive decline and dementia regardless of baseline characteristics: The FINGER trial. Alzheimers Dement. Mar 2018;14(3):263–270. doi:10.1016/j.jalz.2017.09.006

65. Yusuf S, Reddy S, Ounpuu S, Anand S. Global burden of cardiovascular diseases: Part II: variations in cardiovascular disease by specific ethnic groups and geographic regions and prevention strategies. Circulation. Dec 4 2001;104(23):2855–64. doi:10.1161/hc4701.099488

66. Kurian AK, Cardarelli KM. Racial and ethnic differences in cardiovascular disease risk factors: a systematic review. Ethn Dis. Winter 2007;17(1):143–52.

67. Kibria GMA, Crispen R, Chowdhury MAB, Rao N, Stennett C. Disparities in absolute cardiovascular risk, metabolic syndrome, hypertension, and other risk factors by income within racial/ethnic groups among middle-aged and older US people. J Hum Hypertens. Mar 5 2021;doi:10.1038/s41371-021-00513-8

68. Qiu C, Winblad B, Fratiglioni L. The age-dependent relation of blood pressure to cognitive function and dementia. Lancet Neurol. Aug 2005;4(8):487–99. doi:10.1016/S1474-4422(05)70141-1

69. Knopman DS, Roberts R. Vascular risk factors: imaging and neuropathologic correlates. J Alzheimers Dis. 2010;20(3):699–709. doi:10.3233/JAD-2010-091555

70. Tolppanen AM, Solomon A, Soininen H, Kivipelto M. Midlife vascular risk factors and Alzheimer’s disease: evidence from epidemiological studies. J Alzheimers Dis. 2012;32(3):531–40. doi:10.3233/JAD-2012-120802

71. Yaffe K, Vittinghoff E, Hoang T, Matthews K, Golden SH, Zeki Al Hazzouri A. Cardiovascular Risk Factors Across the Life Course and Cognitive Decline: A Pooled Cohort Study. Neurology. Apr 27 2021;96(17):e2212–e2219. doi:10.1212/WNL.0000000000011747

72. Hansson O, Lehmann S, Otto M, Zetterberg H, Lewczuk P. Advantages and disadvantages of the use of the CSF Amyloid beta (Abeta) 42/40 ratio in the diagnosis of Alzheimer’s Disease. Alzheimers Res Ther. Apr 22 2019;11(1):34. doi:10.1186/s13195-019-0485-0

