## Supplementary Methods 1 for "Vascular risk burden is a key player in the early progression of Alzheimer’s disease"

**Supplementary Methods 1. Patient selection.**

1. Analyses of longitudinal cognitive trajectory and risk of clinical progression to cognitive impairment: we included 503 individuals with available baseline medical data and cerebrospinal fluid (CSF) Elecsys biomarkers [amyloid-β_1-42_ (Aβ_1-42_) and tau phosphorylated at threonine 181 (p-tau_181_)]. Also, participants needed to have longitudinal clinical assessments with neuropsychological testing (up to 6 years). Analyses were restricted to subjects with CSF collected within 1.2 years of baseline – here defined as the visit of first neuropsychological assessment.

2. Analysis of longitudinal plasma neurofilament light (NfL): we included 269 participants with available baseline medical data and CSF Elecsys biomarkers (Aβ_1-42_ and p-tau_181_), as well longitudinal NfL measurements (up to 4 years). We also restricted the analysis to individuals who had CSF collected within 1.2 years of baseline – here defined as the visit of first plasma sampling.

3. Analysis of Alzheimer’s disease CSF biomarkers: we included 284 subjects with available baseline medical data and longitudinal CSF Aβ_1-42_ and p-tau_181_ biomarkers measurements (up to 6 years). In this analysis, the time at first CSF collection was defined as baseline.
