## Supplementary Methods 2 for "Vascular risk burden is a key player in the early progression of Alzheimer’s disease"

**Supplementary Methods 2. Vascular risk factor (VRF) burden assessment in the Alzheimer’s Disease Neuroimaging (ADNI) cohort.**

Medical history and use of medications information were manually assessed by three independent authors (JPFS, LAH, and LUDR). The history of the VRFs being assessed [cardiovascular disease, hypertension, diabetes mellitus, hyperlipidemia, stroke or transient ischemic attack (TIA), smoking, atrial fibrillation, and left ventricular hypertrophy]^1, 2^ was considered as the presence or absence of each of them prior or at the baseline visit date. Regarding the use of lipid-lowering, antihypertensive, and antidiabetic drugs, we assessed whether the subject was using or not an included medication (according to the flowchart described in the **Supplementary Fig. 1**) at baseline visit (i.e., current x not current criterion).

The following ADNI tables were assessed, as they presented the detailed information needed for the VRF burden assessment (i.e., complete and specific description of individual diseases, as well as the medications being used):

1. Initial Health Assessment [ADNI3]

2. Recent Medical History Details Log [ADNI1,GO,2]

3. Adverse Events Log [ADNI3]

4. Adverse Events/ Hospitalizations [ADNI1,GO,2]

5. Concurrent Medications Log [ADNI1,GO,2,3]

The authors extracted the information available in these tables and entered the data into a Microsoft Excel spreadsheet. Researchers were blinded to each other’s records. Disagreements were discussed among the three authors who performed the extraction (JPFS, LAH, and LUDR) until a consensus was reached.

In the present study, baseline was defined for each analysis as the first outcome measurement [i.e., plasma NfL, neuropsychological assessment, and cerebrospinal fluid (CSF) Alzheimer’s disease biomarkers]. Given that baseline could differ among analyses (e.g., first plasma NfL measurement could be in a different visit than first neuropsychological assessment), VRF burden was evaluated separately for each set of analyses considering the specific baseline visit. It is important to mention that all variables (e.g., age) were corrected for the baseline date of each particular analysis. Furthermore, CSF measurements used to define Alzheimer’s disease pathophysiology status were the closest to baseline date available and it was restricted to lumbar punctures performed within 1.2 years of the first outcome measurement (i.e., baseline).

Regarding the presence of hyperlipidemia and hypertension, any report suggesting abnormally elevated measurements (e.g., “high blood pressure”, “elevated cholesterol”, "increased cholesterol") was considered as a positive medical history for these conditions. We established this criterion since scrutinizing information related to medication use and laboratory tests (the latter specifically for blood total cholesterol and triglycerides levels) revealed a high agreement between records and patient’s state of disease (e.g., participants described as having “high cholesterol” were usually on lipid-lowering medication use). Noteworthy, no distinctions were made between dyslipidemia and hyperlipidemia.

On the other hand, concerning the presence of diabetes mellitus, we did not consider any report suggesting abnormally elevated measurements of blood glucose (e.g., "hyperglycemia") for a positive medical history of diabetes mellitus. This decision was based on the fact that assessment of data related to medication use and laboratory tests (the latter specifically for blood glucose levels) revealed poor agreement (e.g., participants described as having “hyperglycemia” were usually not on antidiabetic medication use).

When it comes to medications, there were a few cases that did not follow the flowchart described in **Supplementary Fig. 1**:

1. Even though omega-3-acid ethyl esters is a Food and Drug Administration (FDA)-approved medication for lipid-lowering^3^, we did not consider this medication for drug positivity since this term was very likely used to report omega 3 supplements use.

2. Even though Welchol is FDA-approved to be used to improve glycemic control in adults with type 2 diabetes mellitus,^3^ it was not being used for diabetes mellitus treatment in our sample, as the reasons for prescriptions were listed and did not include diabetes. Therefore, the patients taking this drug were not considered positive for antidiabetic medication.

3. Even though drugs of the alpha1-inhibitors class, such as terazosin, doxazosin, prazosin or their corresponding brand names, such as Hytrin and Cardura, are FDA-approved as antihypertensive drugs,^3^ they are not usually used alone for this purpose, as they are one of the last choices in hypertension treatment;^4, 5^ besides, they are many times used for the treatment of benign prostatic hyperplasia.^6^ Therefore, the alpha1-inhibitors drugs were only considered for the presence of hypertension when it was explicitly described that the reason for the prescription was hypertension.

**References**

1.	Bangen KJ, Nation DA, Delano-Wood L, et al. Aggregate effects of vascular risk factors on cerebrovascular changes in autopsy-confirmed Alzheimer's disease. *Alzheimers Dement*. Apr 2015;11(4):394-403 e1. doi:10.1016/j.jalz.2013.12.025

2.	Nation DA, Delano-Wood L, Bangen KJ, et al. Antemortem pulse pressure elevation predicts cerebrovascular disease in autopsy-confirmed Alzheimer's disease. *J Alzheimers Dis*. 2012;30(3):595-603. doi:10.3233/JAD-2012-111697

3.	U.S. Food and Drug Administration. Accessed 30 July 2021, <https://www.accessdata.fda.gov/scripts/cder/daf/>

4.	American College of Cardiology. Accessed 30 July 2021, <https://www.acc.org/latest-in-cardiology/ten-points-to-remember/2017/11/09/11/41/2017-guideline-for-high-blood-pressure-in-adults>

5.	European Society of Cardiology. Accessed 30 July 2021, <https://www.escardio.org/Guidelines/Clinical-Practice-Guidelines/Arterial-Hypertension-Management-of>

6.	American Urological Association. Accessed 30 July 2021, <https://www.auanet.org/guidelines/guidelines/benign-prostatic-hyperplasia-(bph)-guideline/benign-prostatic-hyperplasia-(2010-reviewed-and-validity-confirmed-2014>)
