## Supplementary Table 1 for "Vascular risk burden is a key player in the early progression of Alzheimer’s disease"

**Supplementary Table 1. List of included medications.**

| **Lipid-lowering** | **Antihypertensive** | | | **Antidiabetic** |
| --- | --- | --- | --- | --- |
| Atorvastatin | Accupril | Diltiazem | Microzide | Actos |
| Caduet^a,b^ | Aceon | Diovan | Minoxidil | Avandamet |
| Cholestyramine | Adalat | Diovan HCT^b^ | Monopril | Avandia |
| Colesevelam | Aldactone | Doxazosin | Nadolol | Bydureon |
| Colestid | Altace | Dyazide^b^ | Nifedical | Byetta |
| Colestipol | Amlodipine | Enalapril | Nifedipine | Dulaglutide |
| Crestor | Amlodipine and Benazepril^b^ | Enalapril and Hydrochlorothiazide^b^ | Norvasc | Farxiga |
| Ezetimibe | Atacand | Exforge^b^ | Olmesartan | Glibenclamide |
| Fenofibrate | Atacand HCT^b^ | Felodipine | Perindopril | Glimepiride |
| Gemfibrozil | Atenolol | Fosinopril | Prazosin | Glipizide |
| Lescol | Atenolol and Chlorthalidone^b^ | Furosemide | Procardia | Glucophage |
| Lipitor | Avalide^b^ | Hydralazine | Propranolol | Glyburide |
| Livalo | Avapro | Hydrochlorothiazide | Quinapril | Humalog |
| Lovastatin | Benazepril | Hytrin | Ramipril | Insulin Glargine |
| Lovaza | Benazepril and Hydrochlorothiazide^b^ | Hyzaar^b^ | Spironolactone | Insulin Regular Human |
| Mevacor | Benicar | Indapamide | Telmisartan | Invokamet |
| Niacin | Benicar HCT^b^ | Irbesartan | Terazosin | Janumet^b^ |
| Niaspan | Bisoprolol | Lasix | Tiazac | Januvia |
| Pravachol | Bystolic | Lisinopril | Toprol | Jardiance |
| Pravastatin | Caduet^a,b^ | Lisinopril and Hydrochlorothiazide^b^ | Triamterene | Levemir |
| Repatha | Candesartan | Lopressor | Triamterene and Hydrochlorothiazide^b^ | Lantus |
| Rosuvastatin | Cardizem | Losartan | Twynsta^b^ | Metformin |
| Simvastatin | Cardura | Losartan and Hydrochlorothiazide^b^ | Valsartan | Novolog |
| Tricor | Cartia XT | Lotensin | Vasotec | Onglyza |
| Vytorin^b^ | Carvedilol | Lotrel^b^ | Verapamil | Pioglitazone |
| Welchol | Chlorthalidone | Maxzide^b^ | Verelan | Rosiglitazone |
| Zetia | Clonidine | Metoprolol succinate | Zestoretic^b^ | Toujeo |
| Zocor | Coreg | Metoprolol tartrate | Zestril | Trulicity |
|  | Coversyl plus^b^ | Micardis |  |  |
|  | Cozaar | Micardis HCT^b^ |  |  |

Complete list of included medications (lipid-lowering, antihypertensive, and antidiabetic drugs) detected in the Alzheimer’s Disease Neuroimaging Initiative records for the cognitively unimpaired participants included in the present study.

^a^Caduet is a composition of a lipid-lowering and an antihypertensive drug.

^b^Combination drug (i.e., medication with two or more active ingredients).
