## Supplementary Table 2 for "Vascular risk burden is a key player in the early progression of Alzheimer’s disease"

**Supplementary Table 2. Demographics of the subsample used to assess plasma neurofilament light (NfL) trajectory^a^.**

|  | **(AT)-V-** | **(AT)-V+** | **(AT)+V-** | **(AT)+V+** |
| --- | --- | --- | --- | --- |
| No. | 120 | 110 | 15 | 24 |
| Age at baseline, y | 72.9 (6.2) | 74.0 (6.6) | 74.2 (6.1) | 77.8 (4.8) |
| Male, No. (%) | 47 (39.2) | 61 (55.5) | 7 (46.7) | 9 (37.5) |
| Education, y | 16.5 (2.6) | 16.6 (2.7) | 17.4 (2.5) | 15.1 (2.9) |
| *APOE* ε4 carriers, No. (%) | 23 (19.2) | 33 (30.0) | 9 (60.0) | 11 (45.8) |
| Individual VRFs at baseline, No. (%)^b^ | | | | |
| Cardiovascular disease | 1 (0.8) | 17 (15.5) | 0 (0.0) | 6 (25.0) |
| Hypertension | 26 (21.7) | 96 (87.3) | 5 (33.3) | 23 (95.8) |
| Diabetes mellitus | 1 (0.8) | 23 (20.9) | 0 (0.0) | 0 (0.0) |
| Atrial fibrillation | 1 (0.8) | 9 (8.2) | 0 (0.0) | 2 (8.3) |
| Smoking | 14 (11.7) | 35 (31.8) | 3 (20.0) | 7 (29.2) |
| TIA / stroke | 1 (0.8) | 12 (10.9) | 0 (0.0) | 2 (8.3) |
| Hyperlipidemia | 27 (22.5) | 94 (85.5) | 3 (20.0) | 22 (91.7) |
| VRF burden at baseline | 0.6 (0.5) | 2.6 (0.7) | 0.7 (0.5) | 2.6 (0.6) |
| CSF Aβ_1-42_ at baseline, pg/mL | 1322.0 (414.1) | 1324.7 (415.2) | 689.5 (177.7) | 739.4 (181.5) |
| CSF p-tau_181_ at baseline, pg/mL | 19.2 (7.8) | 20.5 (7.7) | 35.2 (8.5) | 34.0 (8.7) |
| Plasma NfL at baseline, pg/mL | 32.6 (13.8) | 34.4 (15.2) | 39.5 (15.9) | 43.4 (10.8) |
| MMSE score at baseline | 29.2 (1.2) | 28.9 (1.2) | 29.2 (0.9) | 28.6 (1.6) |
| mPACC score at baseline | 0.4 (2.3) | -0.01 (2.9) | 0.5 (2.1) | -1.7 (2.8) |
| Follow-up, y | 2.8 (1.0) | 2.6 (1.0) | 2.6 (1.1) | 2.5 (1.1) |

Participants were stratified according to vascular risk factor (VRF) burden status and Alzheimer’s disease pathophysiology status. Continuous variables are presented as mean (SD). *APOE* ε4 = Apolipoprotein E ε4; Aβ_1-42_ = amyloid-β_1-42_; CSF = cerebrospinal fluid; MMSE = Mini-Mental State Examination; mPACC = modified Preclinical Alzheimer’s Cognitive Composite; p-tau_181_ = tau phosphorylated at threonine 181; TIA = transient ischemic attack.

^a^In this table, baseline refers to the visit of first plasma sampling for NfL measurement.

^b^Prevalence of left ventricular hypertrophy is not displayed in the table because no participant was described to have this condition in the Alzheimer’s Disease Neuroimaging Initiative database.
