## Supplementary Table 3 for "Vascular risk burden is a key player in the early progression of Alzheimer’s disease"

**Supplementary Table 3. Demographics of the subsample used to assess cerebrospinal fluid (CSF) Alzheimer’s disease biomarkers trajectories^a^.**

|  | **V-** | **V+** |
| --- | --- | --- |
| No. | 155 | 129 |
| Age at baseline, y | 73.1 (6.3) | 74.3 (5.7) |
| Male, No. (%) | 59 (38.1) | 70 (54.3) |
| Education, y | 16.7 (2.6) | 16.3 (2.6) |
| *APOE* ε4 carriers, No. (%) | 33 (21.3) | 37 (28.7) |
| Individual VRFs at baseline, No. (%)^b^ | | |
| Cardiovascular disease | 1 (0.6) | 17 (13.2) |
| Hypertension | 33 (21.3) | 105 (81.4) |
| Diabetes mellitus | 0 (0.0) | 19 (14.7) |
| Atrial fibrillation | 1 (0.6) | 7 (5.4) |
| Smoking | 20 (12.9) | 48 (37.2) |
| TIA / stroke | 1 (0.6) | 9 (7.0) |
| Hyperlipidemia | 33 (21.3) | 112 (86.8) |
| VRF burden at baseline | 0.6 (0.5) | 2.5 (0.6) |
| CSF Aβ_1-42_ at baseline, pg/mL | 1252.4 (421.0) | 1169.4 (443.1) |
| CSF p-tau_181_ at baseline, pg/mL | 21.4 (9.0) | 22.2 (9.6) |
| Plasma NfL at baseline, pg/mL^c^ | 32.9 (13.0) | 34.9 (15.0) |
| MMSE score at baseline | 29.2 (1.0) | 29.0 (1.3) |
| mPACC score at baseline | 0.6 (2.3) | -0.5 (2.8) |
| Follow-up, y | 3.2 (1.7) | 3.1 (1.7) |

Participants were stratified according to vascular risk factor (VRF) burden status. Continuous variables are presented as mean (SD). *APOE* ε4 = Apolipoprotein E ε4; Aβ_1-42_ = amyloid-β_1-42_; MMSE = Mini-Mental State Examination; mPACC = modified Preclinical Alzheimer’s Cognitive Composite; NfL = neurofilament light; p-tau_181_ = tau phosphorylated at threonine 181; TIA = transient ischemic attack.

^a^In this table, baseline refers to the visit of first CSF collection for Aβ_1-42_ and p-tau_181_ measurements.

^b^Prevalence of left ventricular hypertrophy is not displayed in the table because only one participant in the V+ group was described to have this condition in the Alzheimer’s Disease Neuroimaging Initiative database.

^c^Assessed in a subset of 140 subjects who had available plasma NfL measurement at the same visit of first CSF collection.
