## Supplementary Table 4 for "Vascular risk burden is a key player in the early progression of Alzheimer’s disease"

**Supplementary Table 4. Crude Hazard Ratios of clinical progression according to vascular risk factor (VRF) burden and Alzheimer’s disease pathophysiology status.**

|  | **Crude Hazard Ratio (95% CI)** | ***P*-value** |
| --- | --- | --- |
| (AT)-V- | 1 (reference) | - |
| (AT)-V+ | 1.3 (0.8 to 2.3) | 0.301 |
| (AT)+V- | 1.2 (0.4 to 3.3) | 0.792 |
| (AT)+V+ | 5.0 (2.5 to 9.8) | < 0.001 |

Cox proportional hazards models evaluating risk of conversion to cognitive impairment (i.e., mild cognitive impairment or dementia) in 6 years according to VRF burden status and Alzheimer’s disease pathology status. Here, model was not fitted with potential confounders. For this analysis, the (AT)-V- group was considered as the reference group. Post-hoc pairwise contrast analysis with Tuckey’s multiple comparison test demonstrated that (AT)+V+ group presented a significantly higher risk of clinical progression in comparison to (AT)-V- (*P* < 0.001) and (AT)-V+ (*P* < 0.001), as well as a marginally greater risk compared to the (AT)+V- (*P* = 0.053). No further differences were observed among the remaining comparisons. CI = confidence interval.
