## Supplementary Figure 1 for "Vascular risk burden is a key player in the early progression of Alzheimer’s disease"

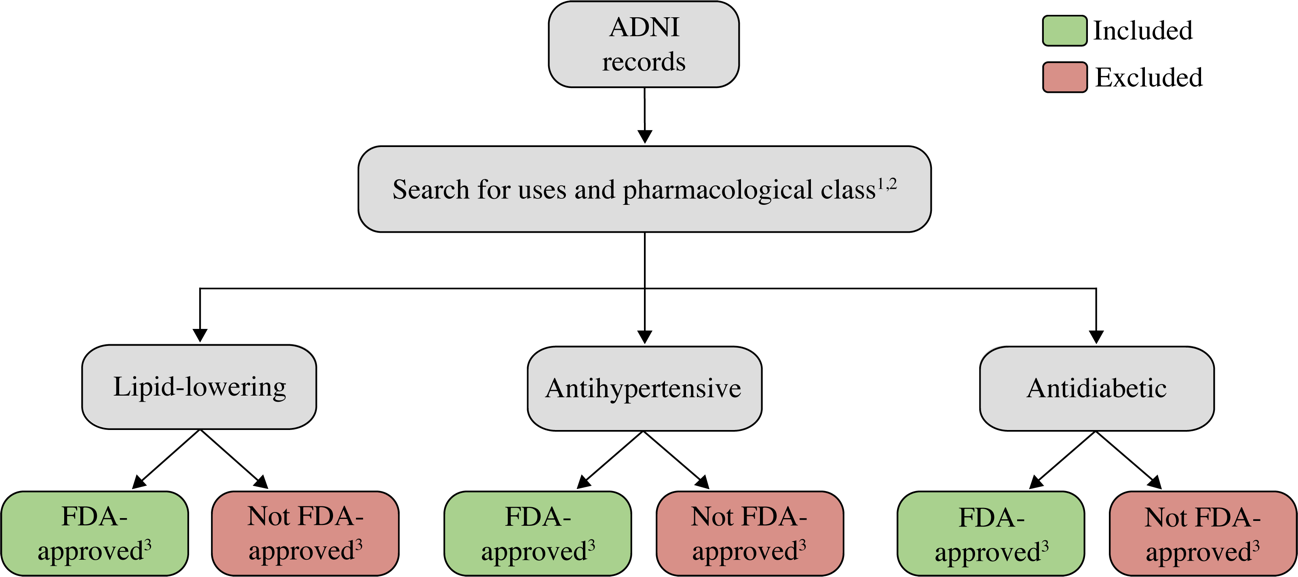

**Supplementary Figure 1. Flowchart of medication assessment.** Each medication described in the Alzheimer’s Disease Neuroimaging Initiative (ADNI) records was searched for uses and pharmacological class,^1, 2^ classified (if applicable) in one of the categories of interest (lipid-lowering, antihypertensive, or antidiabetic), and lastly assessed whether it was, or nor, a Food and Drug Administration (FDA)-approved medication.^3^ The term “Included” refers to drugs that, if being taken at baseline, were considered as a positive history either for hypertension, diabetes mellitus, or hyperlipidemia. The term “Excluded” refers to medications that, even if being taken at baseline, were not considered for a positive history of hypertension, diabetes mellitus, or hyperlipidemia. Exception cases that did not follow this flowchart are described in the previous section (**Supplementary Methods 2**).

**References**

1.	UpToDate. Accessed 30 July 2021, <https://www.wolterskluwer.com/en/solutions/uptodate>

2.	Medscape. Accessed 30 July 2021, <https://www.medscape.com/?src=ppc_google_acq_solo>

3.	U.S. Food and Drug Administration. Accessed 30 July 2021, <https://www.accessdata.fda.gov/scripts/cder/daf/>
